# Epidemiology of leprosy identified through active case detection in six districts of Nepal

**DOI:** 10.1101/2022.08.16.22278814

**Authors:** Ram Kumar Mahato, Uttam Ghimire, Madhav Lamsal, Bijay Bajracharya, Mukesh Poudel, Prashanna Naapit, Krishna Lama, Gokarna Dahal, David TS Hayman, Ajit Kumar Karna, Basudev Pandey, Chuman Lal Das, Krishna Prasad Paudel

## Abstract

**Background:** Nepal has achieved and sustained elimination of leprosy as a public health problem since 2009, but 17 districts and 3 provinces have yet to eliminate the disease. Pediatric cases and grade-2 disabilities (G2D) indicate recent transmission and late diagnosis respectively, which necessitate active and early case detection. This operational research was performed to identify approaches best suited for early case detection, determine community-based leprosy epidemiology, and identify hidden leprosy cases early and respond with prompt treatment.

**Methods:** Active case detection was performed by: house-to-house visits among vulnerable populations (*n=*26,469), contact examination and tracing (*n=*7,608) and screening prison populations (*n=*4,428) in Siraha, Bardiya, Rautahat, Banke, Lalitpur and Kathmandu districts of Nepal.

**Results:** New case detection rates were highest for contact tracing (250), followed by house-to-house visits (102) and prison screening (45) per 100,000 population screened. However, cost per case identified was cheapest for house-to-house visits (Nepalese rupee (NPR) 76,500/case), then contact tracing (NPR90,286/case) and prison screening (NPR298,300/case). House-to-house and contact tracing case paucibacillary/multibacillary (PB:MB) ratios were 59:41 and 68:32; female/male ratios 63:37 and 57:43; pediatric cases 11% in both approaches; and G2D 11% and 5% respectively. Developing leprosy was similar among household and neighbor contacts (Odds ratios (*OR*)=1.4, 95% confidence interval (CI), 0.24-5.85) and for contacts of MB versus PB cases (*OR=*0.7, 0.26-2.0). Attack rates were similar among household contacts of MB cases (0.32%, 0.07-0.94%) and PB cases (0.13%, 0.03-0.73) and neighbor contacts of MB cases (0.23%, 0.1-0.46) and PB cases (0.48%, 0.19-0.98). BCG vaccination with scar presence had a significant protective effect against leprosy (*OR=*0.42, 0.22-0.81).

**Conclusions:** The most effective case identification approach here is contact tracing, followed by house-to-house visits in vulnerable populations and screening in prisons, though house-to-house visits were cheaper. The findings suggest hidden cases, recent transmission, and late diagnosis in the community exist and highlight the importance of early case detection.

## Introduction

Leprosy is a contagious, but low pathogenic and chronic infectious disease caused by *Mycobacterium leprae. Mycobacterium leprae* mainly affects peripheral nerves and skin, which results in progressive physical, psychological and social disability in some cases (1,2). Disability affects the social and working life of infected people; social stigma is a significant consequence of leprosy. The first and prime objective of leprosy control programs is to focus on early case detection so treatment can begin as early as possible after symptoms appear and disability is prevented (3). In 2019, 202,256 new leprosy cases were reported from 118 countries (26.0 per million population), in which 16 countries reported more than 1,000 new cases; the World Health Organization (WHO) South-East Asia Region (SEAR) accounted for 71% of the cases. The new (0.23/10000 population) and child (7.4%) cases, grade 2 disability (G2D, 5.4%) and female (39%) case proportions in 2019 indicate ongoing transmission, late diagnosis and under reported cases in females (4,5).

Nepal has maintained leprosy elimination as a public health problem level at the country level from 2009. However, in 2018, Nepal still reported more than 3,200 cases with a registered prevalence of 0.99/10,000 population. Seventeen districts and 3 provinces have a registered leprosy prevalence of >1/10,000 population, with Madhesh Province (40%) and Lumbini Province (18%) accounting for most cases. The proportion of child, female and G2D cases in Nepal in 2018 were 7.92%, 42% and 4.75% (6). The pediatric cases indicate recent transmission, lower female proportions indicate underreporting, and G2Ds suggest late diagnosis, all threatening the elimination status that Nepal achieved in 2009.

The WHO Global Leprosy Strategy 2016–2020 launched in 2016 envisioned accelerated action towards a leprosy-free world. The indicators for this vision were zero children diagnosed with leprosy and visible deformities, the rate of newly diagnosed leprosy patients with visible deformities <1 per million, and no countries with legislation allowing discrimination on the basis of leprosy. The promotion of voluntary self-reporting is crucial to case detection and for achieving the desired target. The Global Leprosy Strategy 2016–2020 also recommends targeting high-risk and vulnerable groups with increasing active case detection (7). Active case detection is a more effective strategy which enables early diagnosis and treatment and prevents disability and potentially the spread of infection (8,9).

Differing approaches are available for different at-risk populations. House-to-house visits of high-risk and vulnerable populations, such as Dalit, Mushhar and other marginalized communities in Nepal, could identify hidden cases that might transmit the disease in favorable conditions. Contact tracing is a recognized form of undertaking active case detection in a group which is significantly more likely to have leprosy than the general population in high- and low-endemic disease burden countries. Among different types of contacts, household contacts reportedly have 3.5 times greater likelihood of having leprosy than social contacts and almost double that of neighbors; however, even social contacts are 2.5 to 3 times more likely to have leprosy than the general population (10). Studies suggest the most susceptible populations include family contacts of multibacillary (MB) cases, followed by neighboring contacts and the contacts of paucibacillary (PB) cases (11). However, overcrowding within prisons also makes the prison environment conducive for disease spread. Poor diet, lack of hygiene and physical inactivity are enabling factors and hence prisoners are more at risk of transmission compared to general population (12,13). Finally, BCG vaccination, the attenuated Bacillus Calmette-Guérin strain of the related *Mycobacterium bovis* bacteria, also reduces leprosy transmission (14,15).

Here, we use three active case detection methods: 1) house-to-house visits of high-risk and vulnerable populations in Nepali districts with leprosy public health problems; 2) house-to-house visits and examination of contacts of leprosy cases identified between 2-5 years ago; and 3) examination of prisoners to identify early cases in a cross-sectional study. The study also assessed the cost effectiveness of methods to identify active cases and measures of association to highlight key epidemiological features of leprosy in risk areas relevant for control.

## Methods

The general study objective is to determine the epidemiology of leprosy with its protective and risk factors through active and early case detection approach using three active case detection approaches.

### Approach 1

House-to-house visits in communities with high-risk groups and vulnerable populations, such as marginalized habitants of Dalit, Mushhar, and Chamar groups, were undertaken in Rautahat District of Madhesh Province and Banke District of Lumbini Province. In Rautahat, four rural municipalities (Palika): Dewahi Gonahi, Rajpur, Ishnath and Rajdevi were selected in close coordination with district health authorities. These municipalities were considered to have inhabitants from more vulnerable populations. From the four municipalities, 24 sites (wards) covered by 24 health facilities were selected. The same process was followed in Banke, where 27 sites (wards) covered by 27 health facilities from four municipalities, Baijnath, Narainapur, Janaki and Nepalgunj, were selected. A total of 60 to 100 households with inhabitants of marginalized people living in overcrowded houses made of soil or mud, which favored leprosy transmission, were used for the census. Trained local health workers and local female community health volunteers (FCHV) visited the selected sites and performed house-to-house visits, examining all the members present in the household for any signs of leprosy. In total 13,420 and 13,049 individuals were examined respectively in Rautahat and Banke (Table S1). Local trained health workers examined male and FCHV examined female individuals present in the household. Simultaneously, demographic, and epidemiological variables were collected by the trained local health worker.

Members of the households were informed 2 days before the survey and asked to be present at their own household at the time of the survey via the local FCHV. All suspected cases identified by local health workers and FCHV were invited to health facilities and cases confirmed by a dermatologist. After diagnosis confirmation, leprosy cases were treated as per the national protocol.

### Approach 2

Household and neighboring contacts of previously identified confirmed leprosy cases in the previous 2-5 years were examined in Siraha district of Madhesh Province and Bardiya of Lumbini Province by trained local health workers. The cases diagnosed between recent 2-5 years in the respective districts were selected randomly in planning meetings conducted before the implementation of field work. Local trained health workers and FCHVs examined 106 and 177 confirmed leprosy case contacts correspondingly in Siraha and Bardiya. A total of 7608 contacts were screened during the case-contact survey (Table 1).

**Table 1.** Leprosy cases and their contacts screened during a case-contact survey.

| District | Total index cases | Clinical disease | Index cases | Household contacts* | Neighboring contacts** | Total screened population |
| --- | --- | --- | --- | --- | --- | --- |
| Siraha | 106 | MB | 53 | 327 | 1353 | 3170 |
|  |  | PB | 53 | 372 | 118 |  |
| Bardiya | 177 | MB | 107 | 599 | 2102 | 4438 |
|  |  | PB | 70 | 2102 | 1352 |  |
| Total |  |  |  |  |  | 7608 |

\*Household contacts comprised all members >2 years old residing in the index case household.
\*\*Neighboring contacts comprised all individuals >2 years old residing in the nearest 4-6 neighboring houses of index case.

### Approach 3

Siraha (*n=*449), Rautahat (*n=*360), Banke (*n=*826), Bardiya (*n=*319), Lalitpur (*n=*251) and Kathamandu (*n=*2223) prisons were used as screening sites for active case detection using convenience sampling among 4428 prisoners to assess the transmission status of leprosy in prisons. The prisoner population comprised 4229 males and 199 females.

The whole study was carried out between October 2020 and December 2021. Except for the prison population genders, age and gender were only recorded for cases identified.

### Informed consent

In all approaches, participants were requested to give verbal informed consent. As this study was part of regular surveillance of epidemiology and disease control division (EDCD), written informed consent was not taken. Approval for data publication was obtained from EDCD and exemption from ethical review (347/2022) was obtained from ethical board of Nepal Health Research council.

### Statistical analysis

Data collected on paper-based questionnaires, developed by the Leprosy Control and Disability Management Section (LCDMS)/EDCD, were entered in Excel® spreadsheets. Consistency was checked and data analysis done in IBM SPSS statistics 22 and R version 4.2.0. New case detection rate, attack rate (AR) with respect to different demographic variables and types of leprosy cases were calculated with 95% confidence intervals (CI) when appropriate, e.g., using binomial models for the attack rates. Chi-squared tests (χ^2^) and odds ratio (OR) with 95% CIs were calculated for associations between attack rates with respect to different demographic variables and between BCG scar presence and leprosy. To adjust for screened population at risk and index cases present in the case-contact survey, we use simple Poisson regression with an offset for index cases in the population present to assess risk (16), where:

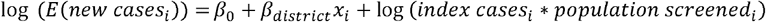

We also simply offset this with population alone, where the offset was log(*population screened*_*i*_) to test the sensitivity of results to this assumption.

### Cost analyses

The costs for each approach were calculated and comprised expenses related to training, orientation, health worker per diems, dermatologist’s fees, expenses for monitoring and supervision and data management. The total cost was divided by total number of patients identified or diagnosed by the approach and derived per unit cost for leprosy case identified. Finally, for discussion, we converted costs from national currencies to US dollars for comparison. We used the date in publications and adjust to 25 December 2021 rates using Google’s default currency convertor provided by Morningstar.

## Results

### Comparison of different approaches of active case detection

New leprosy cases were identified during house-to-house visits (n=27), contact tracing (n=19) and prison screening (n=2) from a total of 38,505 screened people (Table 2). New case detection rates were highest in contact tracing (250 per 100,000 population), followed by house-to-house visits (102 per 100,000) and prison screening (45 per 100,000). However, house-to-house visits were the cheapest cost per case identified at Nepalese rupee (NPR) 76,500/case, followed by contact tracing (NPR 90,286/case) and prison screening (NPR 298,300/case).

**Table 2.** Comparison of different approaches of active case detection.

| Approach | Screened | Suspected cases | Confirmed cases | New case detection rate (/100,000) | PB:MB (%) | F:M (%) | Pediatric cases (%) | G2D (%) | Cost/case (USD\$) |
| --- | --- | --- | --- | --- | --- | --- | --- | --- | --- |
| House-to-house visits | 26469 | 365 | 27 | 102 | 16:11<br>(59:41) | 17:10<br>(63:37) | 3 (11) | 3(11) | 76,500<br>(\$654) |
| Contact tracing | 7608 | 214 | 19 | 250 | 13:6<br>(68:32) | 11:8<br>(57:43) | 2 (11) | 1 (5) | 90,286<br>(\$772) |
| Prison screening | 4428 | 185 | 2 | 45 | 0:2<br>(0:100) | 0:2<br>(0:100) | 0 (0) | 0 (0) | 298,300<br>(\$2550) |

Screened - total numbers screened; PB/MB - paucibacillary/multibacillary ratio; F/M - female/male ratio; G2D – Grade 2 deformity

Just two MB cases were discovered in adult male prisoners. House-to-house and contact tracing case PB:MB ratios were 59:41 and 68:32; female/male ratios 63:37 and 57:43; pediatric cases 11% in both approaches; and G2D 11% and 5% respectively.

### Age and gender of leprosy cases

In aggregate, the number of females among confirmed leprosy cases (27/48) was higher than males, but not significantly different (χ^2^ = 1.3, df = 1, p-value = 0.25) (Figure 1, Table S2).

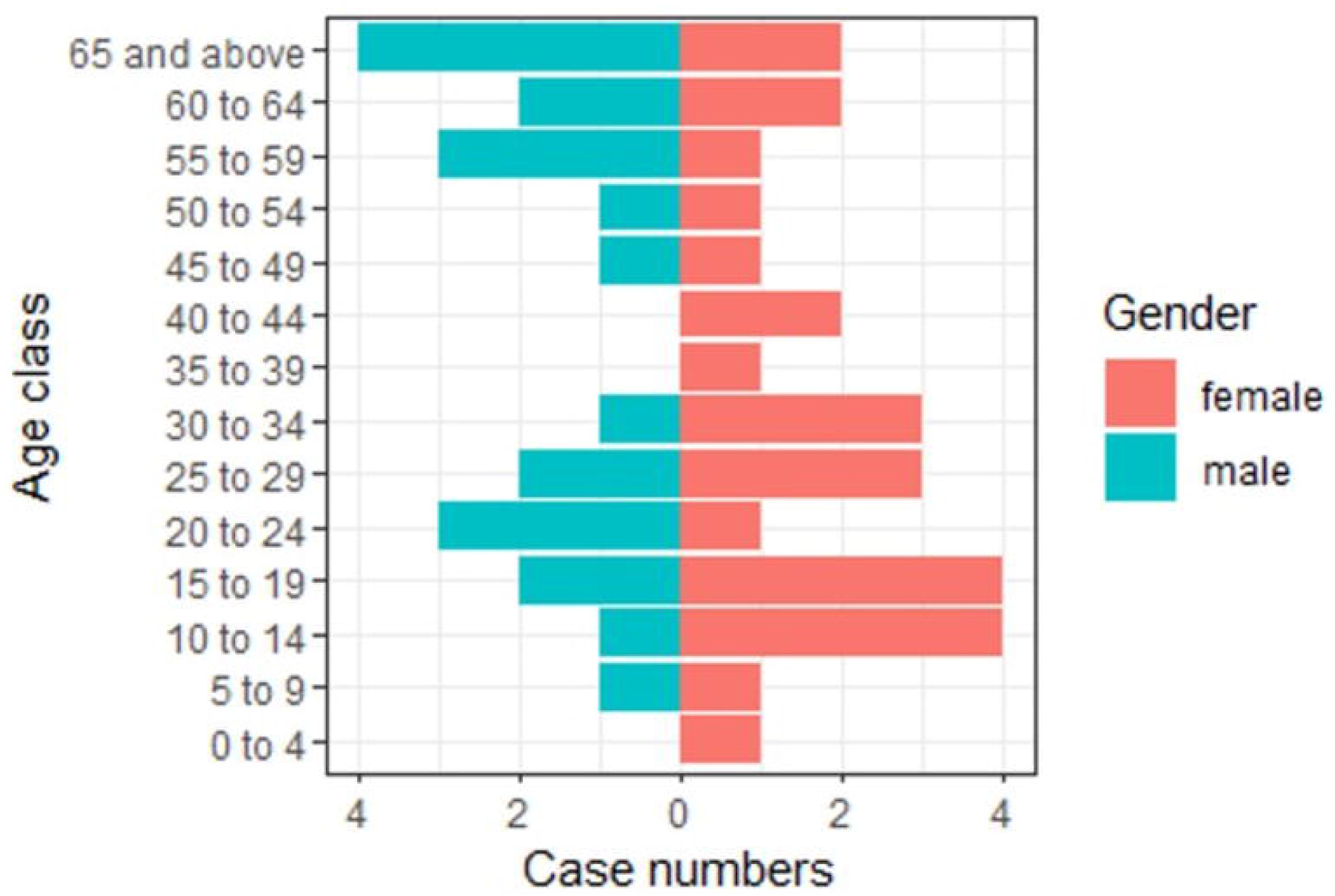

### Attack rate and associations of contacts and leprosy

Individuals with the history of two to five years of proximate contact with leprosy confirmed cases were examined, in which attack rates were higher among household contacts of MB cases (0.32%, 95% CI 0.07-0.94) compared to neighboring contacts of the same cases (0.23%, 0.1-0.46), but this was not significant (χ^2^ = 0.07, df = 1, p-value = 0.9). Neighboring contacts of the PB cases were found to have higher attack rate (0.48%, 0.19-0.98) compared to the household contact (0.13%, 0.03-0.73) of the same cases, but this was again not significant (χ^2^ = 0.8, df = 1, p-value = 0.7) (Table S3, Figure S1).

Differences in related associations, such as household contacts of MB cases being a case compared to neighbours (*OR=*1.4, 95% CI 0.24-5.85) or PB household cases (*OR=*2.5, 95% CI 0.2-129.1) and other variations of these associations were not significantly different. Further data and tables are provided in the supplementary information (Table S4, Figure S2).

The contact tracing of 106 and 177 leprosy index cases in Siraha (*n* = 3170) and Bardiya (*n* = 4438) after 2-5 years of proximate contact with index cases identified 14 new cases Siraha (0.13 cases per index case, 0.07-0.21) and 5 in Bardiya (0.03, 0.01-0.06). This was statistically significant (χ^2^ = 9.8, df = 1, p-value = 0.002), including adjusting for index cases and screened contacts (β = 1.9, standard error = 0.5, p-value < 0.001). The result was insensitive to offsetting to screen population along (β = 1.4, standard error = 0.5, p-value < 0.01).

### Gender, case classification and disability

We found no statistical differences between genders and either having advanced (MB) leprosy (*OR=*0.47, 0.11-1.78, p-value = 0.24) or G2D (*OR=*0.76, 0.05-11.4, p-value = 1) using all 48 new confirmed cases.

### BCG and leprosy transmission

Among the total participants, 569 visited a health facility for confirmation of leprosy and were also inspected for a BCG vaccination scar by a dermatologist. Those participants with the presence of a BCG scar were found to have a significantly lower odds of having leprosy (18 of 341) than without (27 of 201), with an *OR=*0.42 (0.22 – 0.81, p-value = 0.007).

## Discussion

We report 48 new leprosy cases from 38,505 screened people, comprising 29 from house-to-house screening among vulnerable populations, 19 from case-contact tracing and 2 from prisoner screening. House-to-house screening and contact tracing discovered 11% pediatric cases in both approach and 11% and 5% G2D cases respectively, indicating new transmission events and late diagnosis, highlighting the gaps in leprosy control programs.

The new case detection rate was highest in contact tracing (250) followed by house-to-house visits (102) and prison screening (45) in per 100,000 population screened, whereas the most cost-efficient approach here was house-to-house visits (NPR 76,500/case), followed by contact tracing (NPR 90286/case) and prison screening (NPR 298,300/case). These costs per case detected are similar to those reported for other countries, for example case contact tracing was approximately US$529 (vs ∼US$758) in similar study of Nigeria (17), scaled by inflation and using December 2021 exchange rates. The effectiveness and cost efficiency suggest implementing both approaches in parallel may be optimal.

The epidemiological and clinical features of the identified confirmed cases were not significantly different. This is possibly because of the small sample of confirmed cases. The PB:MB ratio differed to the global status (35:65), but greater sample sizes might alter this. Similarly, the female: male ratio differed but with great sample sizes might change to match the national (42:58) and global (40:60) ratios. However, if the findings here are true but simply lacking statistical power due to smaller sample sizes, then these altered ratios could be due to active case detection versus passive case detection and suggest females with leprosy are often hidden with passive surveillance (4). However, details on gender and age were only recorded for cases, so differences in gender and age rates are not available but could be useful for future efforts. Future efforts should also aim for earlier detection through active case detection and to reduce the stigma attached to leprosy, so cases are not hidden.

The transmission attack rates observed in household (0.32%, 95% CI 0.07-0.94) and neighboring (0.13%, 95% CI 0.03-0.73) contacts of MB cases, and household (0.23%, 95% CI 0.1-0.46) and neighboring (0.48%, 95% CI 0.19-0.98) contacts of PB cases were not statistically different. The rates, however, are lower than some other reports, such as 2% in Brazil in 2008 (15). This requires additional studies in more districts to confirm, but the lower attack rate in the current Nepalese situation also indicates progress towards elimination of the disease. The lack of a significant difference in household contacts of cases developing leprosy compared to neighboring contacts (0.78, 95% CI 0.19-2.45) differs from other findings where households contacts may have twice the risk of developing disease compared to neighboring contacts (10). The reasons for this could be sample size and statistical power, or that other factors are either reducing the within-household transmission or increasing the neighbor-case transmission. Again, further work is needed to determine which is occurring, but if reduced within-household transmission, this could be a sign of successful case management. Use of genomic epidemiology my help elucidate transmission chains (18).

The contact tracing of leprosy index cases was conducted after 2-5 years of proximate contact in both Siraha (Madhesh Province) and Bardiya (Lumbini Province), where more new cases per index case in Siraha (14, 0.13 cases per index case, 0.07-0.21) than in Bardia (5, 0.013 per case, 0.01-0.06). The difference was significant, including adjusting for index cases and screened populations. It was reported that leprosy post exposure prophylaxis (LPEP) has been implemented in Bardia for several years.

A further encouraging finding was that BCG vaccination with the presence of a scar has a significant protective effect against leprosy (*OR=*0.42, 95% CI 0.22 – 0.81). The finding is consistent with other findings, such as in Brazil, where the *OR=*0.27 (95% CI 0.13-0.59) (15). The findings suggest BCG immunization programs will successfully contribute to leprosy elimination. For Nepal, this is encouraging, because BCG is given at birth and national coverage is high at 97.8% (95% CI 95.8–98.7) for BCG (19,20). However, like all immunization programs, there are often small pockets of people where there is lower vaccine coverage, and lower BCG is reported for at risk populations such as Madhesi, Dalit, and some religious minorities, who were targeted for screening here (21). Future immunization programs should aim to ensure at risk communities are reached to achieve the leprosy elimination goals.

## Conclusion

The new case detection rates identified in this study suggest sustained levels of transmission in the communities screened. The proportion of pediatric cases (>10%) is evidence of recent transmission and the proportion of G2D confers evidence of late diagnosis and inadequate surveillance in the community. Though not significant, the female: male case ratio being the reverse of the global and national reports from passive case surveillance systems indicates hidden cases in the community, suggesting active surveillance is required to hasten leprosy elimination. The reduced attack rate compared to earlier studies, however, suggest some progress towards disease elimination and BCG vaccination should be given more attention as a tool for elimination, as it reduces transmission. Together, active case detection, through house-to-house visits and contact tracing to detect early and hidden cases, along with the optimal use of BCG, might help block transmission, prevent disabilities, and move Nepal closer towards elimination.

## Contributors

RKM conceived and designed the manuscript; RKM, UG, ML, PN, KL KPP, CLD managed and supervise the data collection; RKM, DTSH analyzed the data; RKM, BB, MKP wrote the original draft KPP, CLD, DTSH, BP, AKK reviewed the manuscript. All authors read, reviewed, and approved the final manuscript.

## Acknowledgements

We are grateful to all the provincial health directors, section chiefs and focal people of the municipalities, health facilities, health offices, where active case detection was carried out for their kind cooperation, direct involvement in case screening, monitoring and supervision. We are more grateful to EDCD LCDMS staff who contributed to coordination and monitoring of the activities. We sincerely express our gratitude to Dr Amrita Shrestha, Consultant Dermatologist of Badegaun PHC, Dr Badri Chapagain, Chief Consultant Dermatologist of Nepalgunj Hospital, Dr Sumit Pandey, Consultant Dermatologist of Nepalgunj Medical college, Dr. Niraj Parajuli, Dr Mahesh Sah, Senior Consultant Dermatologist of Anandban Hospital, Dr Abhisek Bilash Panta, Consultant Dermatologist of Janakpur Hospital and Mr Dipak Bam, Leprosy Expert who worked on the confirmation and validation of suspected cases. This work was supported by grants from WHO through Red book of the Ministry of Health and Population. We are especially thankful to Dr Usha Kiran, coordinator of NTDs section of WHO Nepal who continuously supported us in technical design, and thank Drs Rosemary Barraclough and Jonathan Marshall, Massey University, for helpful comments. DTSH is funded by New Zealand Royal Society Te Apärangi, grant number MAU1701 and a Percival Carmine Chair in Epidemiology and Public Health.

## Data availability

The datasets generated and/or analyzed during the current study are not publicly available due to individual privacy of patients could be compromised but are available from the corresponding author on reasonable request.

## Ethics approval, consent to participate, and consent for publication

The data were generated through contact tracing and house to house visits among vulnerable populations as part of regular program activities by the Epidemiology and Disease Control Division (EDCD). While conducting the field surveillance, consent from the participants was taken verbally in the presence of local health authorities and female community health volunteers (FCHVs). Then all acquired field data were stored at the central database as per the national surveillance system. For this study selected, anonymized data were used and approved for analysis and publication by **Ethical Review Board of Nepal Health Research Council** (**approval number 347/2022 P and approval date: 21 July 2022**) as per the request from EDCD. All data were anonymized for use in writing preparing this manuscript.

## Abbreviations

BCG: Bacille Calmette-Guerin
CI: Confidence Interval
EDCD: Epidemiology and Disease Control Division
FCHV: Female Community Volunteer
G2D: Grade-2 disabilities
LCDMS: Leprosy Control and Disability Management Section
LPEP: Leprosy post exposure prophylaxis
MB: Multibacillary
NPR: Nepalese rupee
OR: Odds ratios
PB: Paucibacillary
SEAR: South-East Asia Region
SPSS: Statistical Package for the Social Sciences
WHO: World Health Organization

