## Supplemental table for "Epidemiology of leprosy identified through active case detection in six districts of Nepal"

**Supplementary Information**

**Tables**

Table S1. Number of households and individuals screened for active case detection

| District | Palika (municipality) | Households screened | People screened |
| --- | --- | --- | --- |
| Rautahat | Dewahi-Gonahi | 400 | 2744 |
|  | Rajpur | 497 | 3238 |
|  | Ishnath | 300 | 1958 |
|  | Rajdevi | 787 | 5480 |
| Banke | Nepalgunj | 891 | 5532 |
|  | Narainapur | 418 | 2229 |
|  | Janaki | 585 | 3612 |
|  | Baijnath | 355 | 1676 |
| Total |  | 4233 | 26469 |

Table S2. Age and sex description of leprosy cases identified during active case detection

| Years | Total cases | Number of female |
| --- | --- | --- |
| 0 to 4 | 1 | 1 |
| 5 to 9 | 2 | 1 |
| 10 to 14 | 5 | 4 |
| 15 to19 | 6 | 4 |
| 20 to 24 | 4 | 1 |
| 25 to 29 | 5 | 3 |
| 30 to 34 | 4 | 3 |
| 35 to 39 | 1 | 1 |
| 40 to 44 | 2 | 2 |
| 45 to 49 | 2 | 1 |
| 50 to 54 | 2 | 1 |
| 55 to 59 | 4 | 1 |
| 60 to 64 | 4 | 2 |
| 60 and above | 6 | 2 |
| Overall | 48 | 27 |

Table S3: Attack rate in contacts of leprosy. See also Figure S1.

| Type of Contact | Cases/number of contacts | Attack rate (%, 95% confidence intervals) |
| --- | --- | --- |
| Household contact of MB cases | 3/926 | 0.32 (0.07-0.94) |
| Household contact of PB cases | 1/757 | 0.13 (0.03-0.73) |
| Neighbouring contact of MB cases | 8/3455 | 0.23 (0.1-0.46) |
| Neighbouring contact of PB cases | 7/1470 | 0.48 (0.19-0.98) |

Table S4. Odds ratios for associations using Fisher's exact test. See also Figure S2.

| Association | Odds ratio (95% CI) | p-value |
| --- | --- | --- |
| MB cases: Household vs neighbour | 1.4 (0.24-5.85) | 0.71 |
| PB cases: Household vs neighbour | 0.28 (0.01-2.17) | 0.28 |
| All cases: Household vs neighbour | 0.78 (0.19-2.45) | 0.8 |
| Household cases: MB vs PB | 2.5 (0.2-129.1) | 0.63 |
| Neighbour cases: MB vs PB | 0.49 (0.15-1.57) | 0.16 |
| All cases: MB vs PB | 0.7 (0.26-2.0) | 0.47 |

Table S5. Impact of BCG on leprosy transmission

| BCG scar | Leprosy | No Leprosy |
| --- | --- | --- |
| Present | 18 | 323 |
| Absent | 27 | 201 |

**Figures**

Figure S1. Attack rates from case-contact surveys. Ninety-five percent confidence intervals (95% CI) are shown.

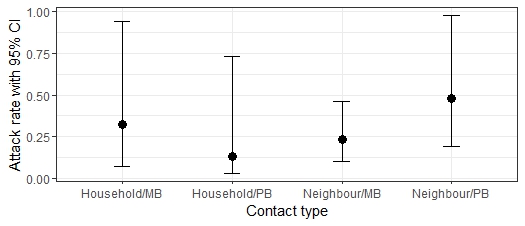

MB – multibacillary; PB – paucibacillary; Household – household contact; Neighbour – neighbouring house contact.

Figure S2. Odds ratios (OR) for associations among cases and contacts. Estimates are shown with 95% confidence intervals (CI). Intervals overlapping *OR=1* are not significantly different. Note the log10 y-axis because of wide confidence intervals.
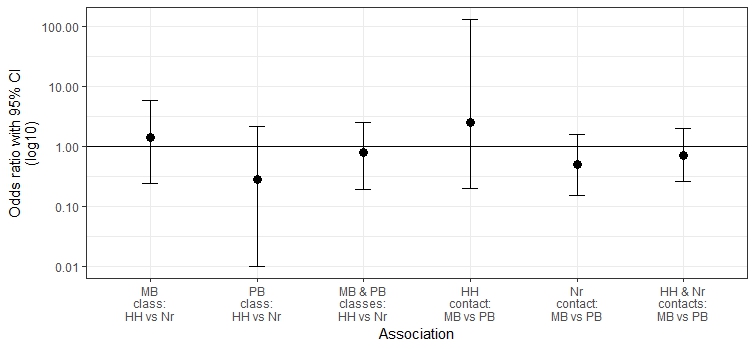

MB – multibacillary; PB – paucibacillary; Household – household contact; Neighbour – neighbouring house contact.
